# Exploring Potential Minocycline–ARH3 Interactions in *ADPRHL2*-Associated CONDSIAS: A Translational Clinical and Computational Study

**DOI:** 10.64898/2026.07.09.26357651

**Authors:** Bita Barazandeh Shirvan, Mojgan Nejabat, Farzin Hadizadeh, Farah Ashrafzadeh, Najmeh Ahangari, Alireza Tavassoli, Henry Houlden, Sajjad Biglari, Mohammad Doosti, Javad Akhondian, Narges Hashemi, Shima Shekari, Mahmoud Mohammadi, Mahmoud Reza Ashrafi, Reza Shervin Badv, Morteza Heidari, Farnoosh Ebrahimzadeh, Zahra Rezaei, Hashem Lashgari Kalat, Zahra Jafari, Elham Pourbakhtiaran, Reza Nejad Shahrokh Abadi, Ehsan Ghayoor Karimiani, Mehran Beiraghi Toosi

## Abstract

**Background:** Stress-induced childhood-onset neurodegeneration with variable ataxia and seizures (CONDSIAS) is a rare autosomal recessive disorder caused by biallelic variants in *ADPRHL2*, which encodes ADP-ribosylhydrolase 3 (ARH3), a key enzyme involved in poly (ADP-ribose) (PAR) metabolism. Although Minocycline has been reported to attenuate PAR-mediated neurotoxicity primarily through modulation of PARP-dependent pathways, whether it may also interact with ARH3 or influence the structural behavior of pathogenic ARH3 variants remains unknown. This study was designed to explore this possibility by integrating clinical observation with computational structural analyses.

**Methods:** Comprehensive clinical evaluation, targeted Sanger sequencing, and in silico pathogenicity analyses were performed. Protein modeling, molecular docking, and 100-ns molecular dynamics simulations were conducted to evaluate the predicted structural consequences of the p.Thr79Pro variant and to explore potential interactions between ARH3 and Minocycline.

**Results:** A homozygous *ADPRHL2* variant (NM_017825.3:c.235A>C; p.Thr79Pro) was identified in a child with CONDSIAS. Computational analyses predicted reduced structural stability and increased conformational flexibility of the mutant ARH3 protein relative to the wild-type structure. MM-GBSA calculations estimated differences in binding free energies between the wild-type (−34.51 kcal/mol) and mutant (−39.76 kcal/mol) ARH3-Minocycline complexes, suggesting subtle differences in their predicted energetic profiles. Clinically, neurological progression appeared stable, with improved motor function observed during approximately one year of follow-up and no notable treatment-related adverse effects.

**Conclusions:** By integrating clinical observations with computational structural analyses, this study provides preliminary computational support for the hypothesis that Minocycline may influence ARH3 conformational behavior in addition to its proposed effects on PARP-dependent pathways. Although these findings do not demonstrate direct molecular binding or therapeutic efficacy, they provide a biologically plausible framework for future biochemical, cellular, and functional investigations.

## 1. Introduction

Stress-induced childhood-onset neurodegeneration with variable ataxia and seizures (CONDSIAS; MIM 618170) is a rare autosomal recessive neurodegenerative disorder caused by biallelic pathogenic variants in *ADPRHL2*, which was first described in 2018 (1). Patients with CONDSIAS exhibit a broad and heterogeneous clinical spectrum, including ataxia, epilepsy, gait instability, nystagmus, hearing loss, ptosis, ophthalmoplegia, dysarthria, muscle weakness, axonal neuropathy, myopathy, dysmetria, and tongue fasciculations (2–4). Early neurodevelopment is typically normal, followed by stress-triggered neurological deterioration that may progress rapidly and result in severe disability or early childhood mortality (5). ADP-ribosylhydrolase 3 (ARH3), encoded by *ADPRHL2* on chromosome 1p34, is a key regulator of the ADP-ribosylation (ADPr) pathway through its role in poly (ADP-ribose) (PAR) degradation (5). The coordinated balance between PAR synthesis by poly (ADP-ribose) polymerases (PARPs) and PAR degradation by ARH3 is essential for maintaining appropriate cellular stress responses (6). In response to oxidative stress, excitotoxicity, and other cellular insults, PARPs synthesize PAR polymers that activate downstream stress-response pathways (7). ARH3 counteracts this process by hydrolyzing PAR and PAR-derived products. Loss of ARH3 function disrupts PAR homeostasis, resulting in pathological PAR accumulation and increased susceptibility to stress-induced neurodegeneration (8,9). Evidence from model organisms further supports the pathogenic importance of PAR dysregulation. Neuron-specific knockdown of *Parg* in flies, a model of impaired PAR degradation, results in severe neurodegeneration and lethality that can be partially rescued by pharmacological PARP inhibition, including treatment with Minocycline (3). These findings highlight PAR-related pathways as potential therapeutic targets in CONDSIAS. However, whether the reported biological effects of Minocycline are mediated solely through PARP-related mechanisms remains unknown. In particular, it is unclear whether Minocycline may also influence ARH3 through direct or indirect molecular interactions, or whether pathogenic ARH3 variants alter its predicted interaction profile. In the present study, we integrated clinical assessment with protein structural modeling, molecular docking, and molecular dynamics simulations to explore potential interactions between Minocycline and ARH3 in CONDSIAS. Rather than attempting to demonstrate direct molecular binding or therapeutic efficacy, our objective was to generate mechanistic hypotheses regarding whether Minocycline might influence the structural behavior of mutant ARH3 and thereby provide a rationale for future biochemical and functional investigations.

## 2. Materials and Methods

### 2.1 Clinical Evaluation and Ethical Approval

This study employed an integrated clinical and computational approach to investigate the potential molecular and clinical implications of Minocycline in a patient with CONDSIAS. Ethical approval was obtained from the Ethics Committee of Mashhad University of Medical Sciences, Mashhad, Iran (IR.MUMS.MEDICAL.REC.1404.249). Written informed consent for publication was obtained from the patient’s legal guardians. The study was conducted in accordance with the STROBE (Strengthening the Reporting of Observational Studies in Epidemiology) reporting guideline, and the completed checklist is provided in the Supplementary Material.

### 2.2 Genetic Analysis

Given the presence of a similarly affected sibling and a previously identified familial variant, targeted Sanger sequencing was performed to confirm the suspected *ADPRHL2* variant. Primers targeting exon 2 of *ADPRHL2* were designed using the Primer3 web-based tool (Forward: 5′-AGGTGAGCAGGAGGCTCTCATC-3′; Reverse: 5′-GATGCCTCTAAGGGTTGGGAGAG-3′). The target region was amplified by polymerase chain reaction (PCR), and sequencing was performed using capillary electrophoresis on an ABI 3500xl DNA Analyzer (Applied Biosystems, Foster City, CA, USA). Variant pathogenicity was further evaluated using in silico prediction tools, including MutationTaster, PolyPhen-2, CADD, and SIFT.

### 2.3 In Silico Structural and Dynamic Analyses

#### 2.3.1 Conservation and Structural Modeling

Evolutionary conservation at the nucleotide level was assessed using the UCSC Genome Browser. The experimentally determined X-ray crystal structure of ARH3 (PDB ID: 6D36) was used as the template for structural analyses. The mutant model was generated using ColabFold v1.6.1. Structural quality was assessed using the UCLA-DOE LAB SAVES v6.0 server. Residue-specific evolutionary conservation was evaluated using the ConSurf server (10).

#### 2.3.2 Physicochemical Properties and Stability Predictions

The Physicochemical properties of ARH3, including molecular weight, amino acid composition, theoretical isoelectric point (pI), and instability index, were calculated using ProtParam. The predicted effects of the substitution on protein stability were evaluated using I-Mutant 2.0 and MUpro, both of which employ machine-learning models trained on experimentally validated datasets (11,12). Predicted structural consequences of the variant, including alterations in residue size, charge, hydrophobicity, and local conformation, were further assessed using the HOPE server (13).

#### 2.3.3 Molecular Docking and Molecular Dynamics Simulations

The three-dimensional (3D) structure of Minocycline (PubChem CID: 54675783) was retrieved from the PubChem database for molecular docking studies. Ligand and protein preparation were performed using Vega ZZ and AutoDock Tools, including the addition of polar hydrogen atoms and removal of crystallographic water molecules. Molecular docking was carried out using AutoDock Vina with a grid box of 22.5 × 22.5 × 22.5 Å centered at coordinates (x = −2.33, y = 1.028, z = −0.001). The highest-ranked docking poses were selected for subsequent protein– ligand interaction analyses. To investigate the dynamic behavior of Minocycline in complex with both wild-type and p.Thr79Pro ARH3 proteins under explicit solvent conditions, molecular dynamics (MD) simulations were performed using GROMACS 2022 with the CHARMM27 force field. Ligand parameters were generated using the SwissParam web server. The systems were solvated, neutralized by adding Na□ and Cl□ ions, and subjected to energy minimization to eliminate steric clashes. Equilibration was subsequently performed under constant volume (NVT) and constant pressure (NPT) conditions. Production MD simulations were conducted at 300 K and 1 atm using a 2-fs integration time step for 100 ns. Trajectory analyses were performed using standard GROMACS utilities. Protein conformational stability and dynamics were evaluated by calculating the root-mean-square deviation (RMSD), root-mean-square fluctuation (RMSF), and radius of gyration (Rg).

## 3. Results

### 3.1 Clinical and Genetic Findings

The proband was a male child born to consanguineous parents after an unremarkable prenatal and perinatal course. An affected sibling had developed progressive gait ataxia, speech regression, and recurrent seizures during early childhood and subsequently died due to disease-related complications (Figure 1A). Early developmental milestones were achieved appropriately before disease onset, which was characterized by recurrent febrile seizures followed by progressive gait ataxia and motor regression. Brain magnetic resonance imaging demonstrated mild cerebellar vermian atrophy (Figure 1B). Neurological examination during disease progression revealed generalized hypotonia, joint hyperlaxity ،inability to walk independently, and markedly limited expressive language. Comprehensive metabolic workup (serum ammonia, lactate, thyroid and liver panels; venous blood gas; serum amino acids; MS/MS; urine organic acids) was normal. Clinical whole-exome sequencing performed in the affected sibling identified a homozygous *ADPRHL2* variant (NM_017825.3: c.235A>C). Targeted Sanger sequencing subsequently confirmed the same homozygous variant in the proband, while both parents were heterozygous carriers, consistent with autosomal recessive inheritance (Figure 1C). The variant met multiple ACMG/AMP pathogenicity criteria, including PS1, PM2, PP3, and PP1. Population frequency data and in silico pathogenicity predictions are summarized in Table 1.

**Figure 1.**
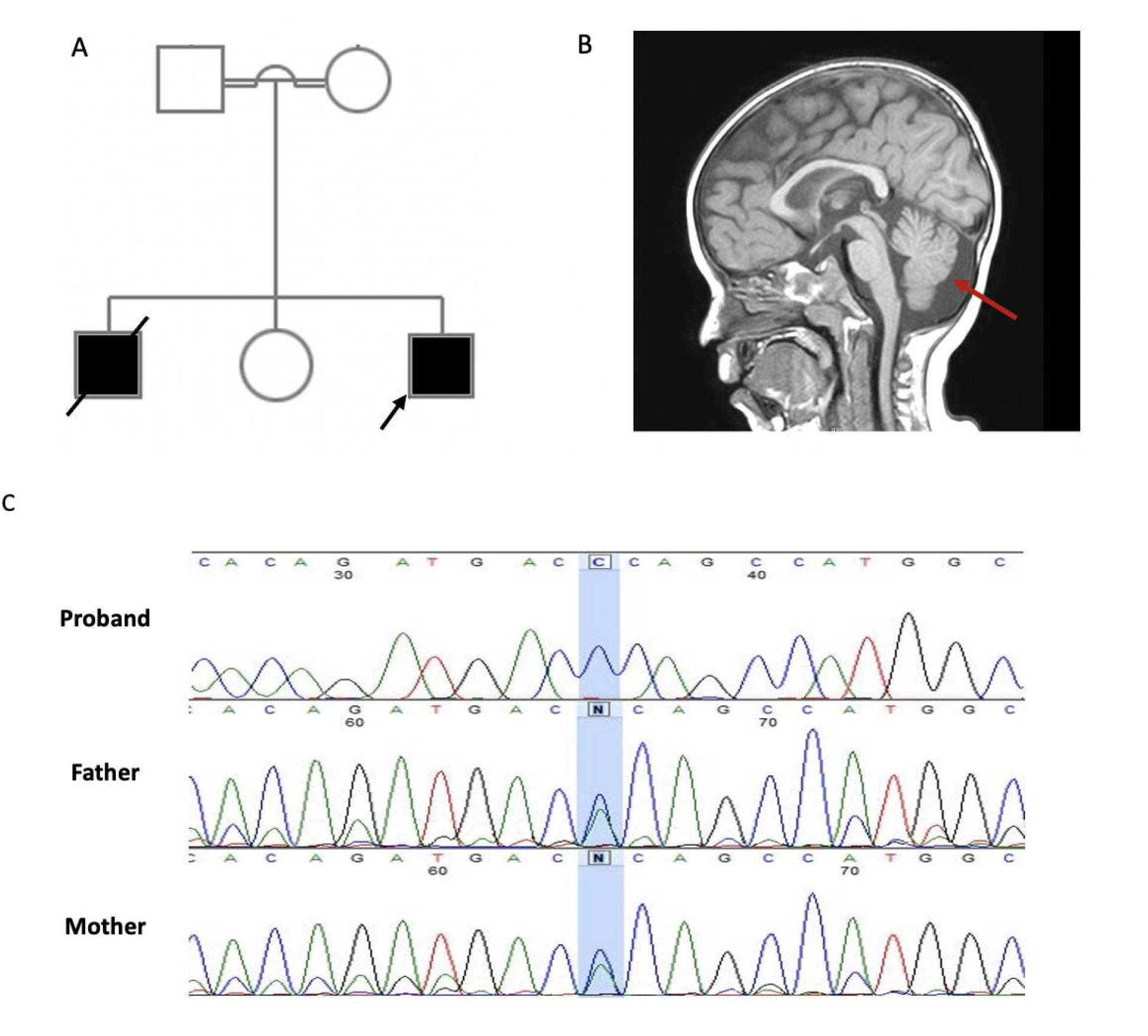
(A) Pedigree of a consanguineous family harboring the *ADPRHL2* c.235A>C variant*. Filled symbols indicate affected individuals; double lines indicate consanguinity.* (B) Sagittal T1-weighted brain MRI of the proband showing mild cerebellar vermian atrophy (red arrow). (C) Sanger sequencing chromatograms for each family member. The proband is homozygous for the c.235A>C variant; both parents are heterozygous carriers; the unaffected sibling lacks the variant.

**Table 1.**
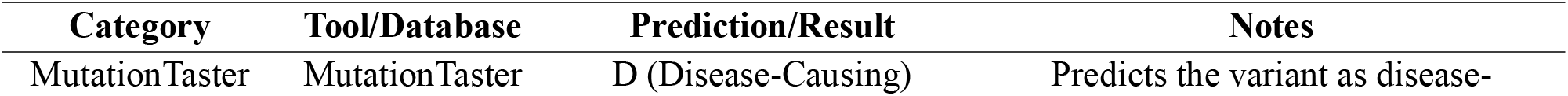
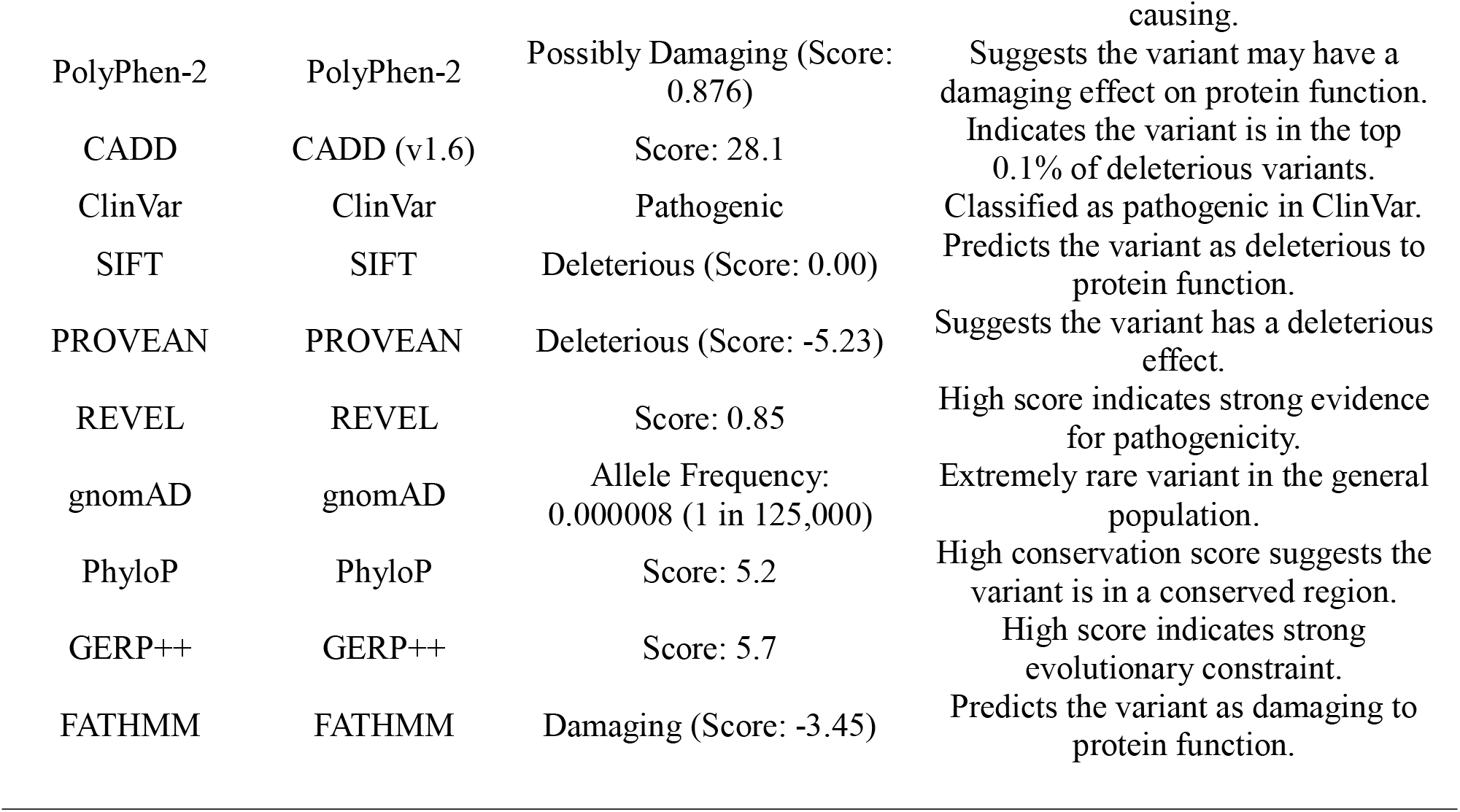
*In silico* pathogenicity predictions for the *ADPRHL2* variant (NM_017825.3: c.235A>C; p.T79P) across multiple bioinformatics tools, summarizing its predicted impact on protein function and clinical significance.

Based on previous experimental evidence implicating PAR-related pathways in CONDSIAS models, off-label Minocycline treatment (100 mg/day) was initiated. Concomitant antiepileptic therapy with valproate (10 mg/kg/day) and clobazam (10 mg/day) was continued without modification. During approximately one year of longitudinal follow-up, modest clinical improvement was observed in motor function. The patient became able to stand with physiotherapy assistance, and expressive language progressed to the production of simple two-word phrases. Gross motor function improved from Gross Motor Function Classification System (GMFCS) level IV to level III, although expressive language plateaued. No further neurological deterioration was observed during the follow-up period, seizure control remained stable, and no clinically significant treatment-related adverse effects were documented.

### 3.2 Evolutionary Conservation and Structural Modeling

ConSurf analysis indicated that the p.Thr79Pro substitution occurs at a highly conserved ARH3 residue under strong evolutionary constraint, suggesting potential functional relevance of this position. Phylogenetic analysis of homologous sequences further demonstrated conservation of the corresponding genomic region (chr1:36,091,262–36,091,272) across multiple species. The genomic organization of *ADPRHL2*, ARH3 domain architecture, and residue-level conservation scores are presented in Figure 2A–C. Three-dimensional structural models of the wild-type and p.Thr79Pro ARH3 proteins are shown in Figure 3A–C. Structural quality assessment using the UCLA-DOE LAB-SAVES v6.0 server yielded an ERRAT score of 93.97, indicating acceptable model quality and non-bonded atomic interactions. VERIFY3D analysis demonstrated that 95.59% of residues achieved a 3D–1D compatibility score ≥0.1, supporting the reliability of the predicted models for subsequent computational analyses.

**Figure 2.**
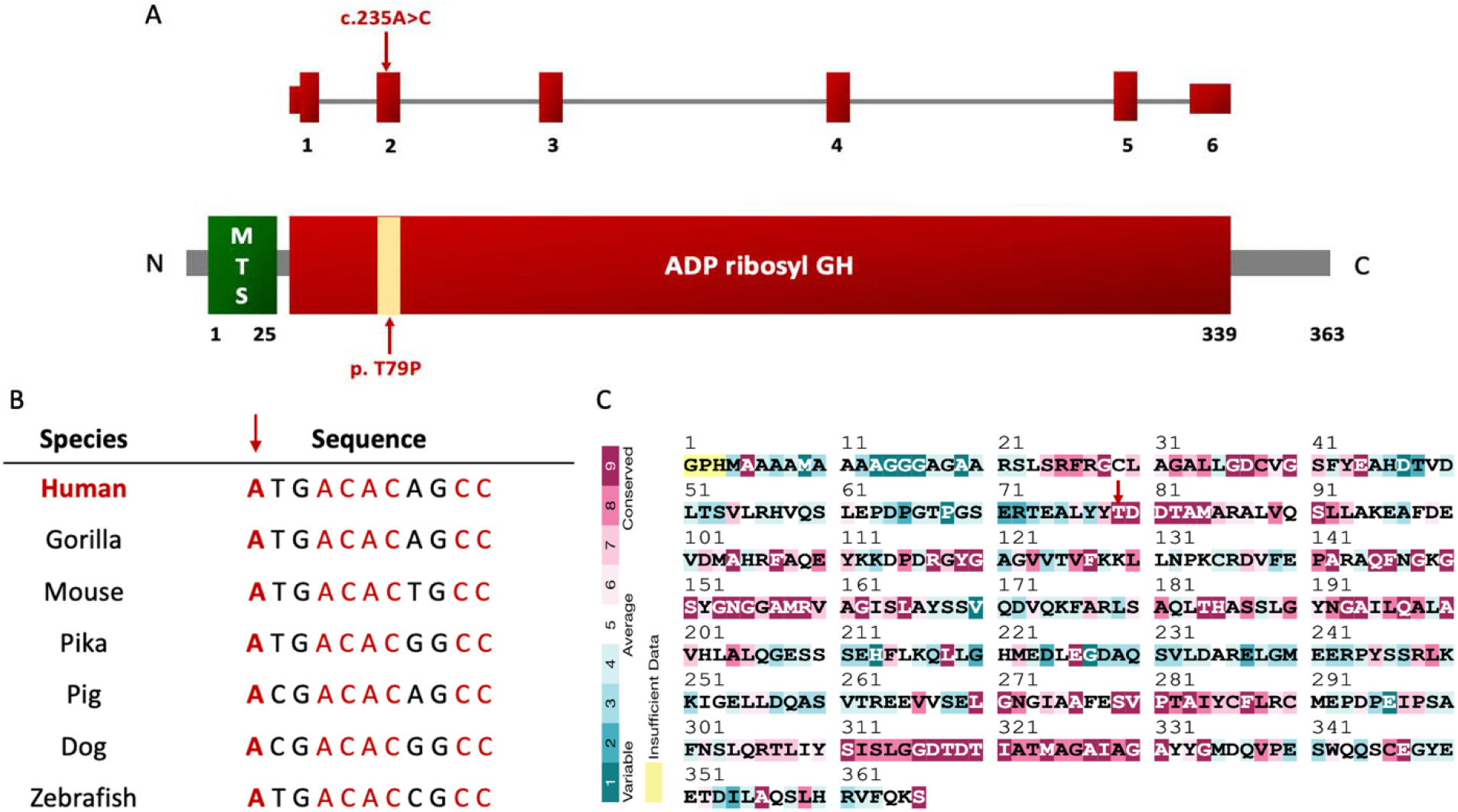
Structural and computational characterization of the ARH3 p. Thr79Pro variant. (A) The variant location within the ADPRHL2 gene and ARH3 protein. (B) Multiple sequence alignment across orthologous species shows that the variant occurs within a highly conserved region, with conserved residues highlighted in red. (C) Amino acid conservation scores assessed through the ConSurf server. Scores range from 1 (variable) to 9 (highly conserved. The arrow marks the mutated residue (Thr79), located within a highly conserved region.

**Figure 3.**
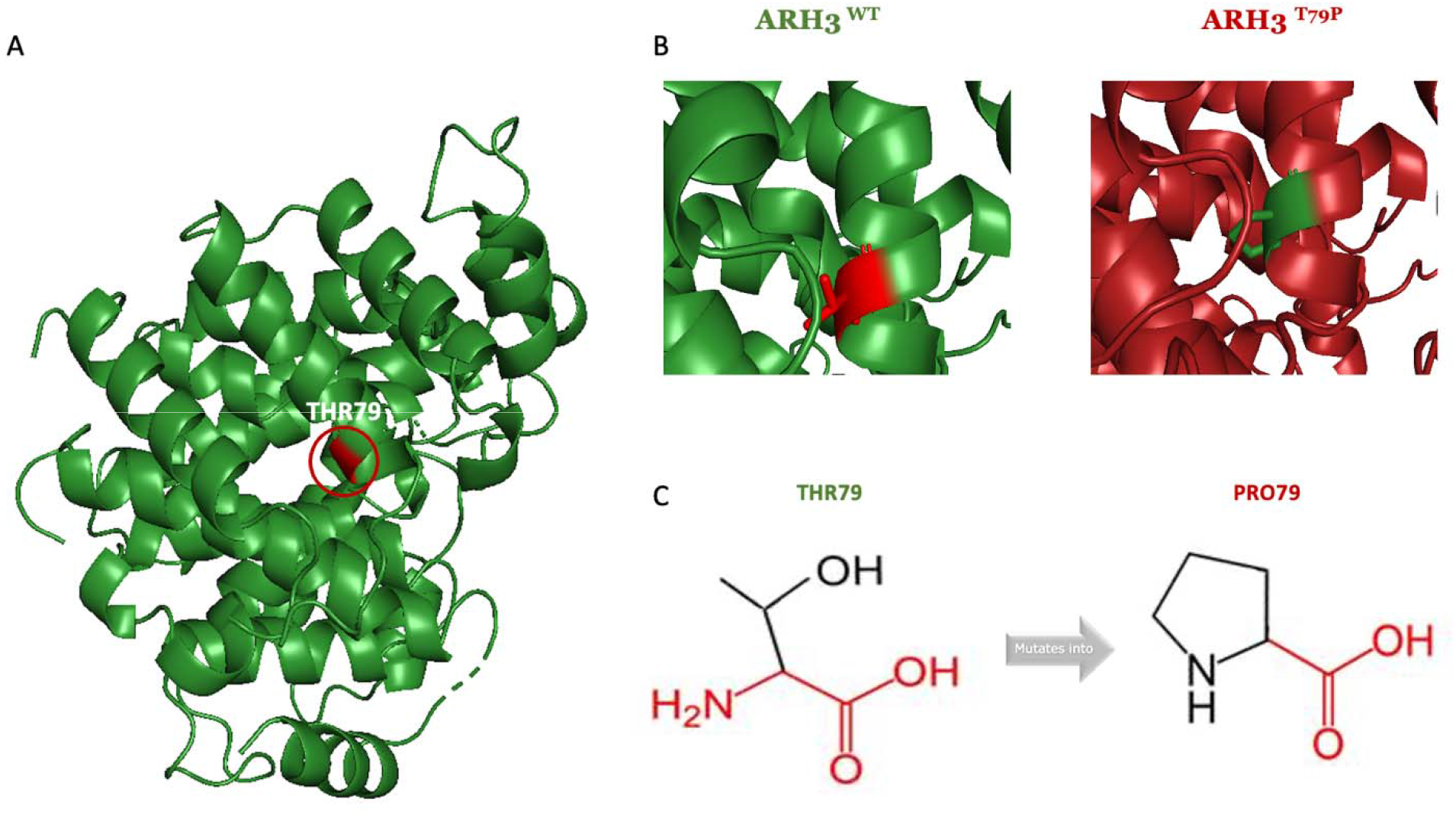
Structural modeling and comparative analysis of wild-type and mutant ARH3 proteins. (A) Three-dimensional structure of the ARH3 protein highlighting the site of the amino acid substitution (Thr79). (B) Superimposed models of wild-type ARH3 (green) and the T79P mutant (red), showing conformational differences induced by the substitution. (C) Detailed structural comparison of the wild-type (left) and mutant (right) residues at position 79. Backbone atom (shared by all amino acids) are shown in red, and side chain atoms (unique to each residue) are shown in black.

### 3.3 Physicochemical Properties and Predicted Structural Stability

ProtParam analysis demonstrated minimal differences in the predicted physicochemical properties of wild-type and p.Thr79Pro ARH3 proteins. The predicted molecular weights were 38,946.75 Da for wild-type ARH3 and 38,942.76 Da for the mutant protein, with instability index values of 33.10 and 34.04, respectively. Both proteins exhibited an acidic theoretical isoelectric point (pI = 4.95), while GRAVY values remained similar (−0.161 for wild type and −0.164 for mutant), indicating minimal predicted changes in overall physicochemical characteristics.In contrast, multiple computational algorithms predicted a potential destabilizing effect of the p.Thr79Pro substitution. MUpro predicted reduced protein stability with a ΔΔG value of −0.89 kcal/mol, supported by both support vector machine and neural network models. Similarly, I-Mutant 2.0 predicted decreased stability (ΔΔG = −0.43 kcal/mol at pH 7.0 and 25°C). HOPE analysis predicted local structural alterations associated with the substitution, including increased hydrophobicity and potential disruption of a hydrogen bond involving Asp34 within the protein core. MetaRNN predicted a high probability of functional impact (score = 0.90). Collectively, these independent computational predictions suggest that the p.Thr79Pro variant may negatively influence ARH3 structural stability; however, experimental validation is required to confirm these effects.

### 3.4 Molecular Docking and Molecular Dynamics Simulations

Molecular docking analyses predicted comparable Minocycline interaction profiles for wild-type and p.Thr79Pro ARH3 structures. AutoDock Vina scores were −7.5 kcal/mol for wild-type ARH3 and −7.6 kcal/mol for the p.Thr79Pro variant. Predicted MM-GBSA binding free energies were −34.51 kcal/mol and −39.76 kcal/mol, respectively. In both predicted complexes, Minocycline was predicted to interact with conserved ARH3 residues, including Tyr116, Gly117, Gly150, His182, and Asp314 (Figure 4). No major differences in predicted ligand orientation were observed between wild-type and mutant ARH3 structures, suggesting that the p.Thr79Pro substitution does not markedly alter the predicted Minocycline interaction interface. To further investigate the effect of the variant on protein conformational behavior, 100-ns molecular dynamics simulations were performed for apo and Minocycline-bound wild-type and mutant ARH3 systems. RMSD trajectories demonstrated stable conformational behavior in all simulated systems, with apparent stabilization after approximately 20 ns (Figures 5 and 6A–B). Mean RMSD values were 0.29 ± 0.06 nm for apo wild-type ARH3 and 0.35 ± 0.05 nm for apo mutant ARH3. In Minocycline-bound systems, RMSD values were 0.29 ± 0.05 nm for wild-type ARH3 and 0.33 ± 0.04 nm for the p.Thr79Pro Rg analysis revealed minimal differences in global protein compactness between wild-type and mutant ARH3. Apo systems showed mean Rg values of 1.96 ± 0.02 nm for wild-type ARH3 and 1.97 ± 0.01 nm for the mutant protein. Similarly, Minocycline-bound complexes exhibited comparable Rg values of 2.005 ± 0.02 nm and 2.004 ± 0.01 nm, respectively (Figure 6C–D). RMSF analysis revealed similar residue-level fluctuations in both apo and complex forms. Peaks were observed at residues 1, 66, 131, and 363, located in the N-terminal, loop region, and C-terminal (which showed the highest flexibility) (Figure 7A–C). The p.Thr79Pro variant showed modest increases in local conformational fluctuations without evidence of major global structural rearrangements. A summary of molecular dynamics parameters is provided in Table 2. Overall, these computational analyses indicate that the p.Thr79Pro substitution may influence ARH3 structural stability and conformational dynamics while preserving a similar predicted Minocycline interaction profile. These findings do not demonstrate direct molecular binding or functional modulation of ARH3 by Minocycline but provide a structural framework supporting further biochemical, biophysical, and cellular investigations.

**Figure 4.**
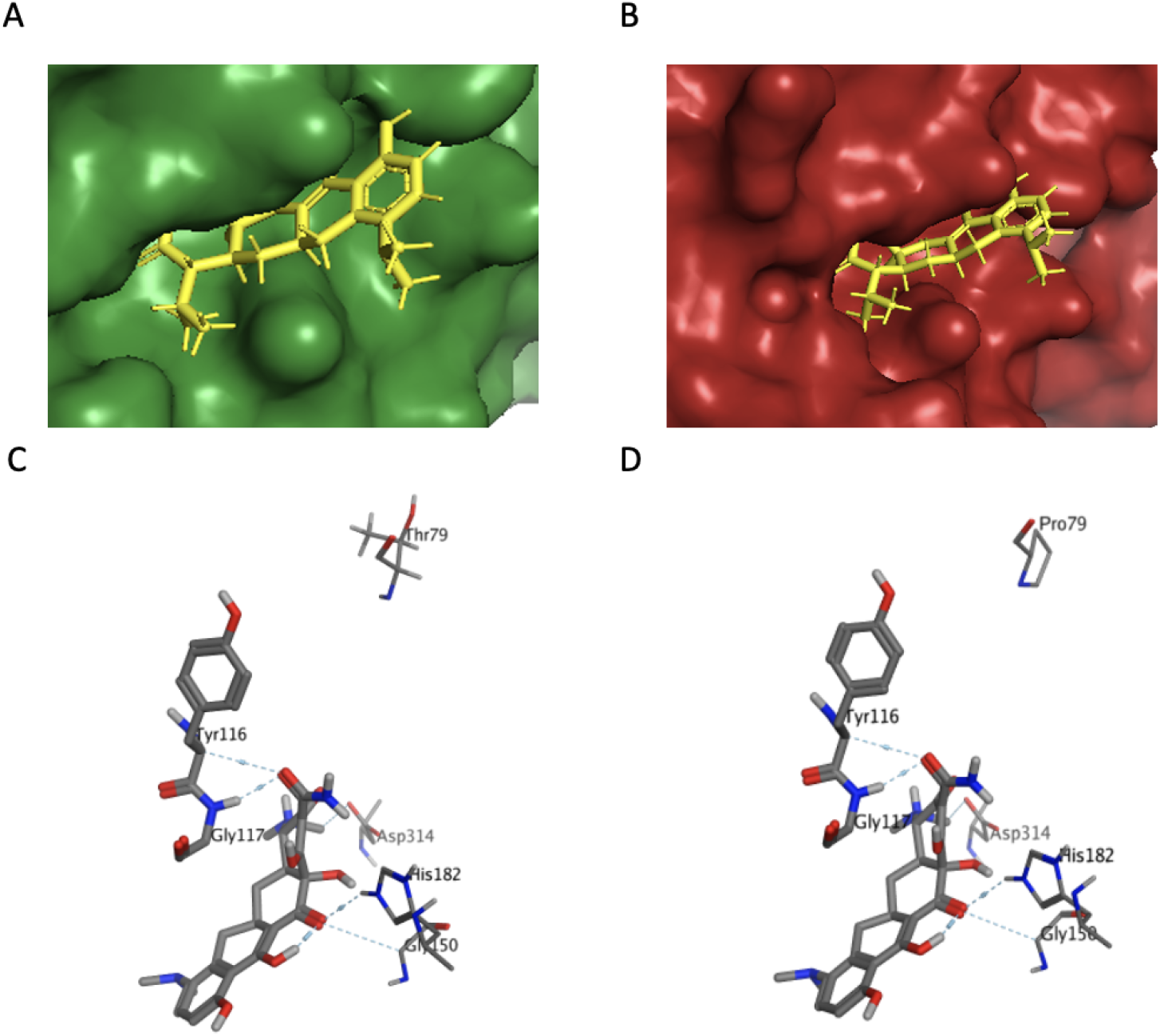
Molecular docking analysis of wild-type and mutant ARH3 with minocycline. (A, B) Surface representations of the docking complexes showing wild-type ARH3 (green) and mutant ARH3 (red) bound to minocycline (yellow sticks). (C, D) Two-dimensional interaction diagram of wild-type (C) and mutant (D) ARH3 with minocycline, highlighting hydrogen-bonding residues. The positions of Thr79 (wild type) and Pro79 (mutant) are indicated.

**Figure 5.**
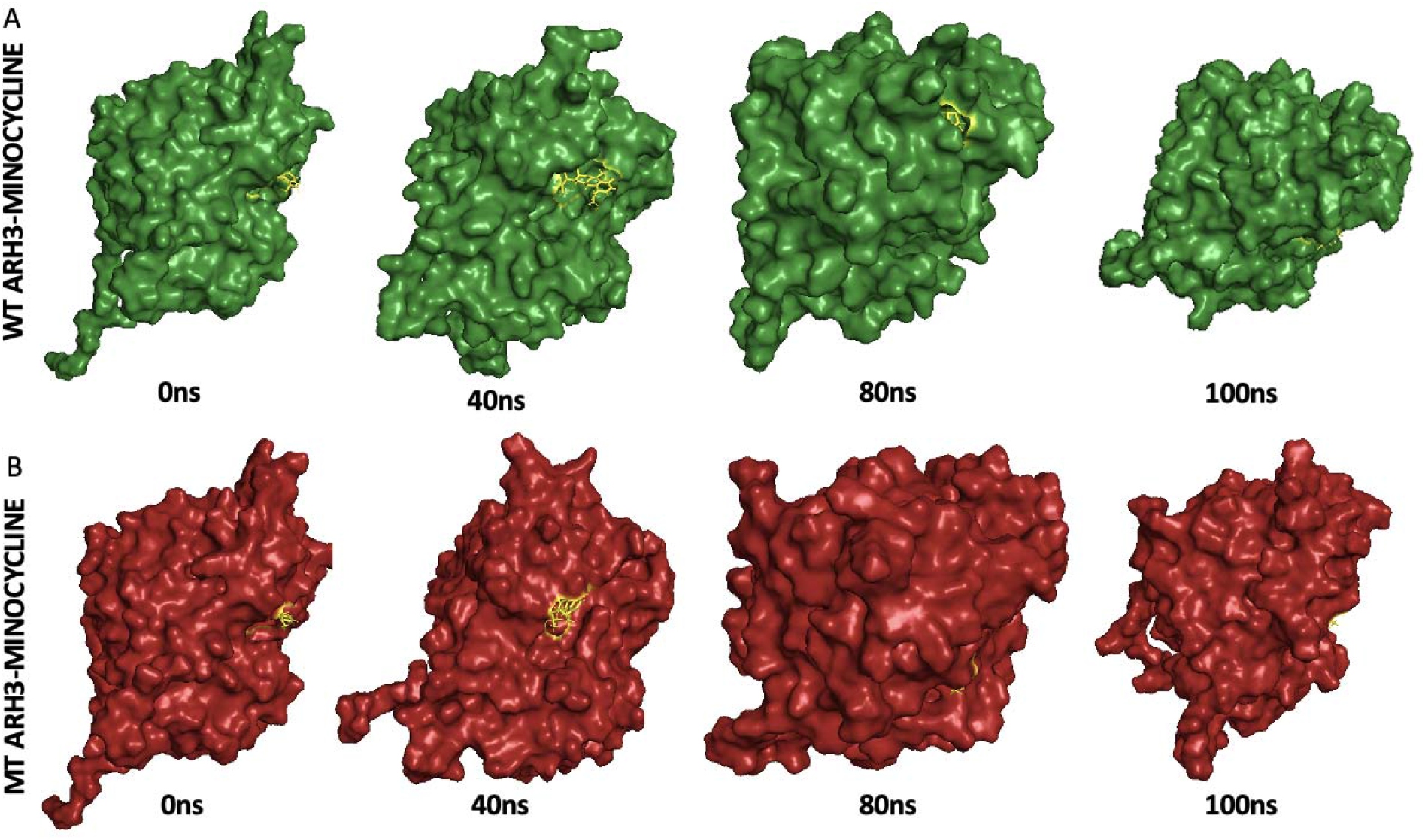
Molecular dynamics (MD) simulation snapshots of ARH3–minocycline complexes at 0 ns, 40 ns, 80 ns, and 100 ns. (A) Wild-type (WT) ARH3–minocycline complex. (B) Mutant-type (MT) ARH3–minocycline complex.

**Figure 6.**
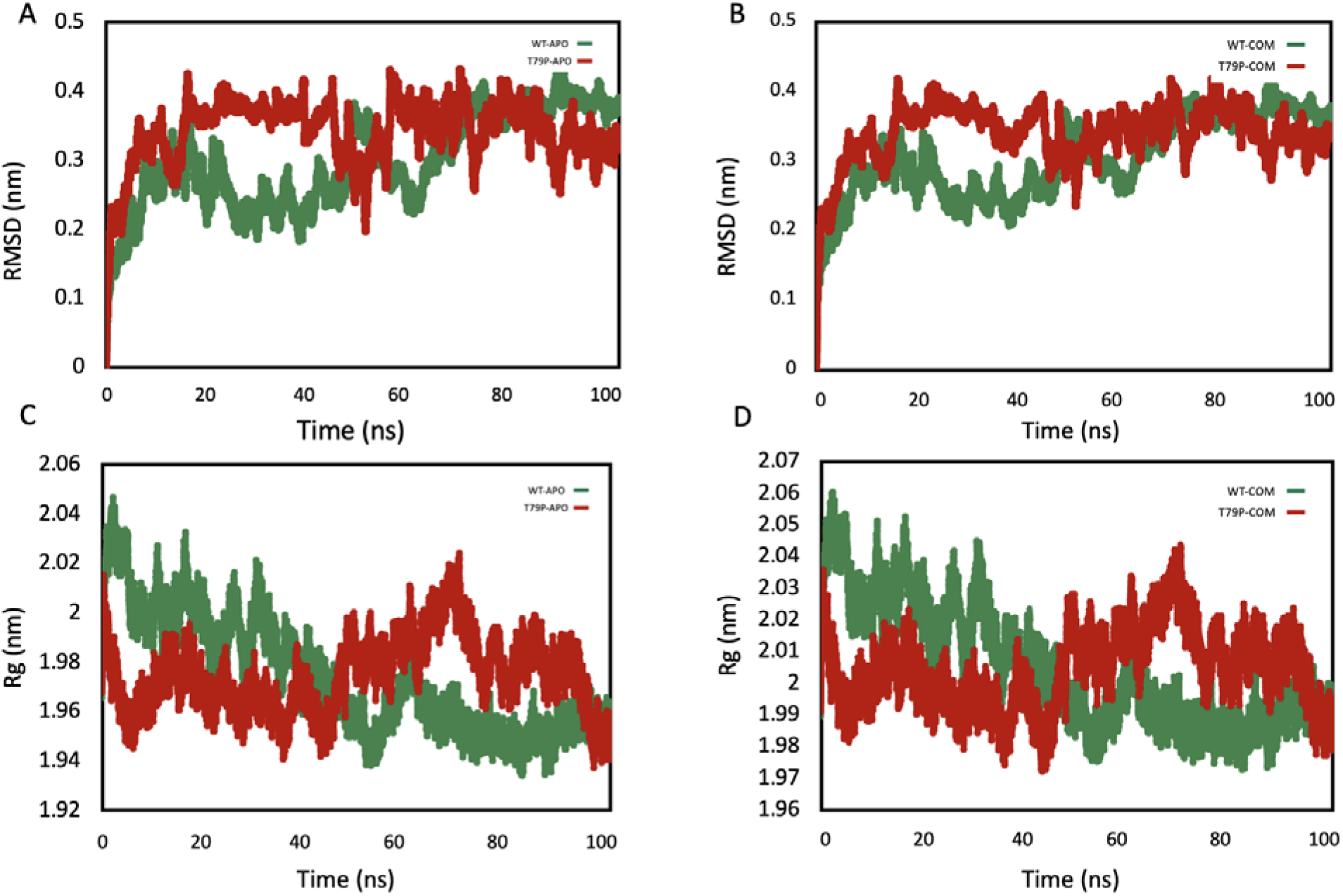
Structural stability analysis of wild-type and mutant ARH3 using RMSD and radius of gyration (Rg) plots. (A) RMSD plot of wild-type (WT) and mutant (T79P) ARH3 in the apo state. (B) RMSD plot of WT and mutant ARH3 in complex with minocycline. (C) Radius of gyration (Rg) plot of WT and mutant ARH3 in the apo state. (D) Rg plot of WT and mutant ARH3 in complex with minocycline.

**Figure 7.**
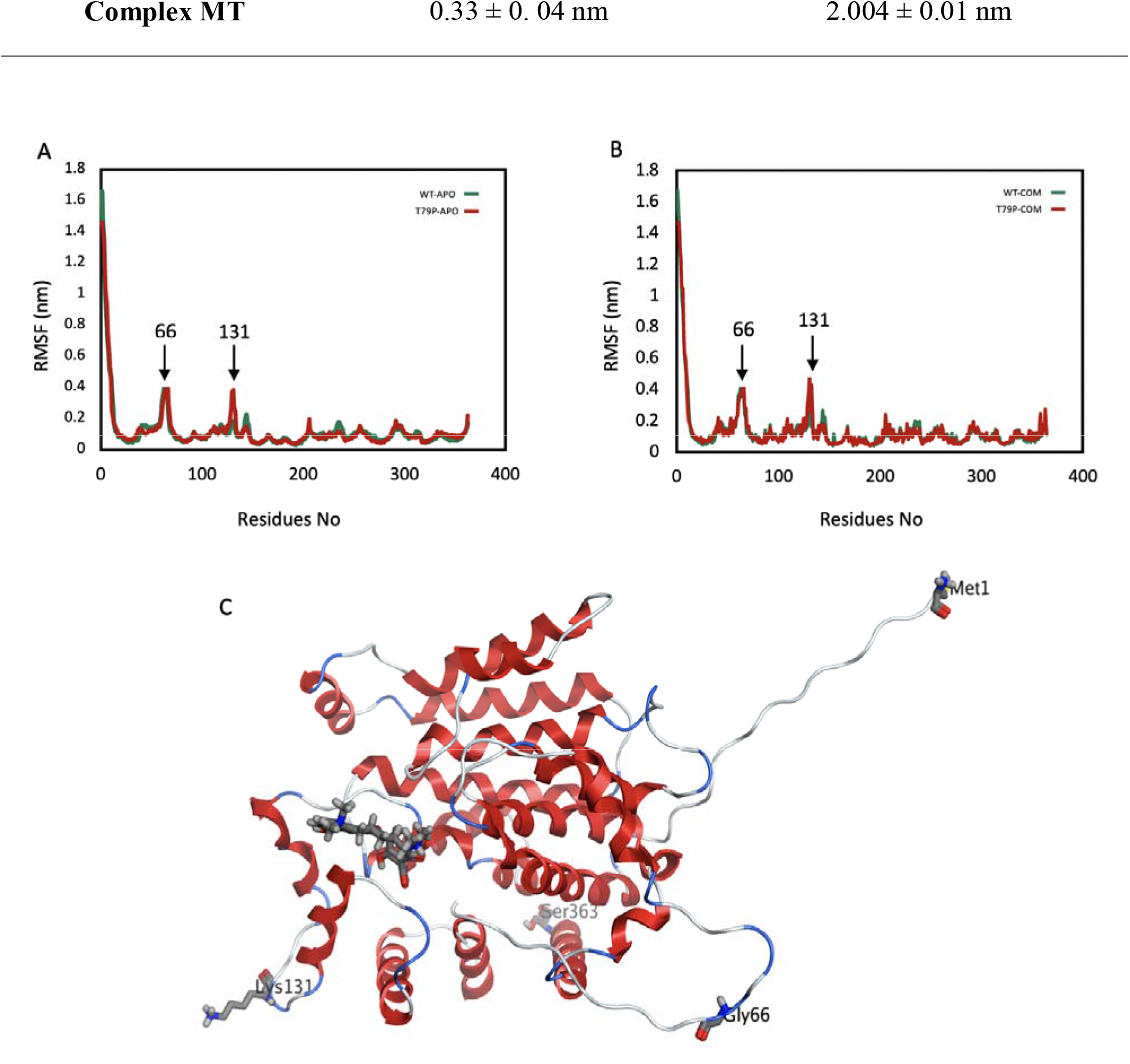
Residue flexibility analysis of wild-type and mutant ARH3 using RMSF plots. (A) RMSF plot of wild-type (WT) and mutant (T79P) ARH3 in the apo state. (B) RMSF plot of WT and mutant ARH3 in complex with minocycline. Peaks were observed at residues 1, 66, and 131. (C) Structural representation showing the positions of minocycline and residues 1 (C-terminal), 66, 131, and 363 (N-terminal) with high RMSF values.

**Table 2.**
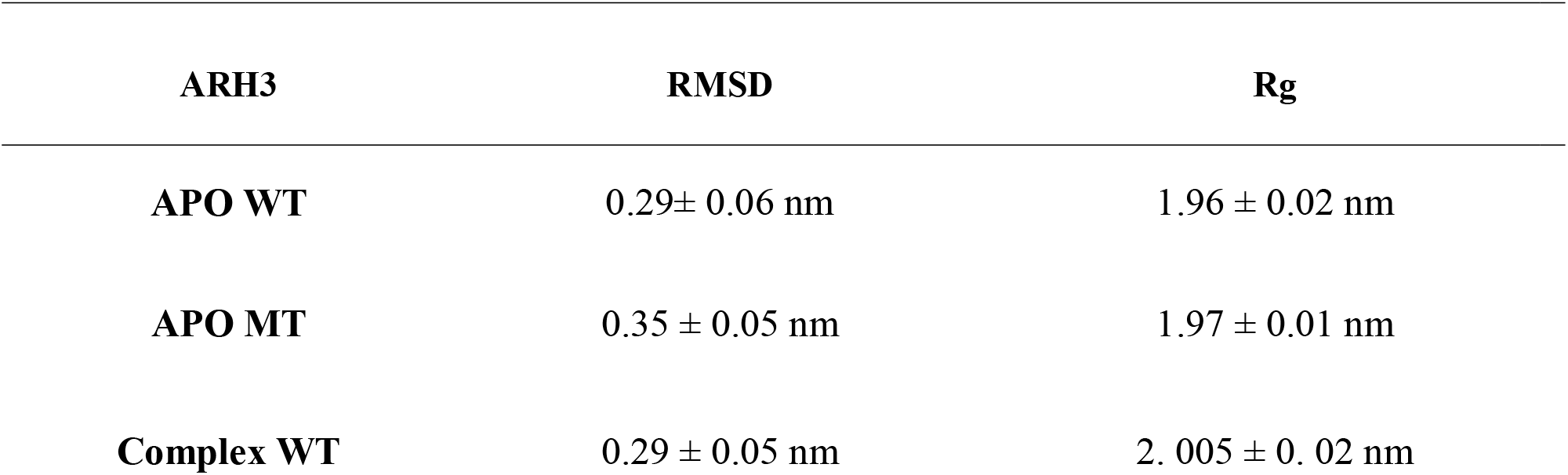
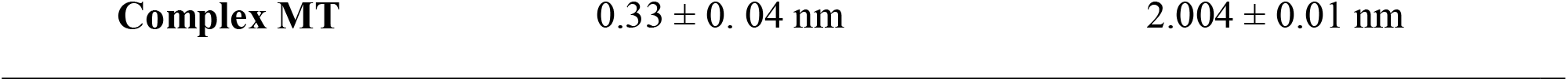
The RMSD and Rg results for WT and mutant ARH3 (T79P) in apo state and complex with Minocycline.

## 4. Discussion

Pathogenic variants in *ADPRHL2* cause CONDSIAS, a rare neurodegenerative disorder characterized by progressive neurological deterioration. ARH3, encoded by *ADPRHL2,* is a key regulator of poly (ADP-ribose) (PAR) metabolism and plays an essential role in maintaining ADP-ribosylation homeostasis (14,15). Under physiological conditions, PAR polymers synthesized by PARP enzymes during cellular stress are degraded through the coordinated actions of PARG and ARH3, thereby limiting excessive PAR accumulation and PAR-mediated neuronal injury (16–18). Loss of ARH3 function disrupts this balance and increases neuronal vulnerability to stress. In flies, neuron-specific knockdown of *Parg*, a functional paralog of *ADPRHL2*, results in severe neurodegeneration and lethality that can be partially rescued by pharmacological PARP inhibition, including Minocycline treatment (3). Furthermore, modulation of ADP-ribosylation pathways has been investigated in several studies and diseases, including cancer, infectious diseases, cardiovascular disorders, and neuromuscular diseases (14,19–22). A previous case report also described clinical improvement following Doxycycline administration in a patient with an *ADPRHL2* variant, although the underlying mechanism remained unclear (23). Anecdotal reports have also suggested possible clinical benefit from high-dose vitamin supplementation (24), but these observations have not been systematically evaluated. Despite these observations, whether Minocycline exerts biological effects beyond PARP modulation, including potential interactions with ARH3 itself, has not previously been explored. The clinical observation that motivated this study was the stabilization of neurological progression accompanied by modest clinical improvement following initiation of off-label Minocycline therapy. Although these findings are compatible with the proposed PARP-mediated mechanism of Minocycline, they prompted us to consider whether an additional mechanism might contribute to the observed clinical course. Specifically, we hypothesized that Minocycline could interact with mutant ARH3 and potentially influence its structural behavior. To explore this possibility, we performed molecular docking and molecular dynamics simulations as a hypothesis-generating approach. Our computational analyses consistently predicted that the p.Thr79Pro substitution affects a highly conserved residue and reduces ARH3 structural stability. These predictions are consistent with previous experimental studies demonstrating reduced ARH3 abundance, decreased thermal stability, and altered secondary structure in patient-derived fibroblasts carrying this variant (3). Molecular docking predicted comparable Minocycline interaction profiles in wild-type and mutant ARH3, with similar docking scores and preservation of interactions involving several conserved residues. Although MM-GBSA calculations suggested differences in predicted binding free energies, the mutation was not predicted to abolish the potential interaction interface. Molecular dynamics simulations further indicated modestly increased conformational flexibility of the mutant protein while preserving its overall structural integrity. Collectively, these findings suggest that Minocycline may influence the conformational landscape of mutant ARH3; however, whether such effects occur under physiological conditions remains unknown and requires experimental validation. Although the observed clinical stabilization is encouraging, this report describes a single patient who also received concomitant antiepileptic therapy and physiotherapy. Therefore, a causal relationship between Minocycline administration and the observed clinical course cannot be established, and alternative explanations, including disease heterogeneity and supportive rehabilitation, cannot be excluded. The principal contribution of this study is the integration of a clinical observation with computational structural analyses to generate a biologically plausible and experimentally testable hypothesis regarding potential Minocycline–ARH3 interactions in CONDSIAS. While our findings do not establish direct ligand binding, protein stabilization, or therapeutic efficacy, they provide a structural framework for future biochemical, cellular, and in vivo studies aimed at determining whether ARH3 represents an additional molecular context contributing to the biological effects of Minocycline. Several limitations should be acknowledged. First, this study is based on a single patient and therefore cannot establish treatment efficacy or generalizability. Second, all structural analyses relied on computationally derived models and molecular simulations rather than experimentally determined protein structures. Finally, molecular docking, MM-GBSA calculations, and molecular dynamics simulations generate structural hypotheses but cannot establish direct ligand binding or functional rescue in the absence of biochemical validation. Accordingly, additional experimental studies will be required to determine whether Minocycline directly influences ARH3 stability, enzymatic activity, or PAR metabolism under physiological conditions.

## 5. Conclusion

This study integrates a clinical observation with computational structural analyses to propose a testable hypothesis regarding potential Minocycline–ARH3 interactions in CONDSIAS. Although these findings do not demonstrate direct molecular binding or therapeutic efficacy, they suggest that the pathogenic p.Thr79Pro variant is predicted to retain a similar interaction profile with Minocycline despite reduced computationally predicted structural stability. These observations provide a structural rationale for future biochemical, cellular, and in vivo investigations to determine whether ARH3 represents an additional molecular context that may influence the biological effects of Minocycline in *ADPRHL2*-associated neurodegeneration.

## Supporting information

Supplementary- check list

## Declarations

### Ethics approval and consent to participate

Ethical approval was obtained from the Ethics Committee of Mashhad University of Medical Sciences, Mashhad, Iran (Approval Number: IR.MUMS.MEDICAL.REC.1404.249). Written informed consent was obtained from the patient’s parents before participation. The study was registered in the Iranian Registry of Clinical Trials (IRCT) under registration number (IRCT20250624066242N7), and the study protocol was conducted in accordance with the approved registration details.

### Consent for publication

Written informed consent was obtained from the patient’s parents before participation to publish the acquired results.

### Author contribution(s)

B.B.S. in-silico analysis and writing the draft, M.N. in-silico analysis, F.H. data interpretation and manuscript revision, F. A. clinical examination, N.A. writing the draft, data collection and data interpretation, A.T. data interpretation and manuscript revision, H.H. data interpretation and manuscript revision, S. B. data interpretation, manuscript revision, M. D. data interpretation and manuscript revision, J. A. clinical examination, N.H. clinical examination and manuscript revision, Sh. Sh., M. M., M. R. A., R. Sh., M. H., F. E., Z. R. H., F.E., L.K. and E. P. manuscript revision, Z. J. software, E. G. K. data interpretation, manuscript revision, M. B. T. conception, organization, clinical examination, data interpretation, manuscript revision, R.N.S.A writing the draft, manuscript revision. All authors have read and approved the final version of the manuscript, were responsible for the decision to submit the manuscript for publication, and agree to be accountable for all aspects of the work in ensuring that questions related to the accuracy or integrity of any part of the work are appropriately investigated and resolved. M. B. T and E. G. K. accessed and verified the underlying data.

## Acknowledgements

The authors appreciate the Clinical Research Development Unit of Ghaem Hospital, Mashhad University of Medical Sciences, Mashhad, Iran, for their valuable support and assistance in this study, by providing the opportunity to the authors and helping to improve the quality of the manuscript. The authors would also like to thank Sina Beiraghi Toosi for his contribution to the language editing and refinement of the manuscript.

## Availability of data and materials

All the data and materials used are included in the manuscript. The datasets used and/or analyzed during the current study are available from the corresponding author upon reasonable request.

## Use of AI-Assisted Tools

AI-assisted tools (ChatGPT, OpenAI) were used solely to support language refinement and to improve the clarity, grammar, and readability of the manuscript. All scientific content, including study design, data interpretation, computational analyses, molecular modeling, molecular dynamics simulations, and formulation of conclusions, was generated, reviewed, and verified by the authors. No AI tools were used to generate, modify, or manipulate research data, images, molecular structures, or analytical outputs. The authors take full responsibility for the accuracy, integrity, and originality of all scientific content presented in this work.

