## Supplementary- check list for "Exploring Potential Minocycline–ARH3 Interactions in *ADPRHL2*-Associated CONDSIAS: A Translational Clinical and Computational Study"

The STROBE reporting checklist

For checking that observational epidemiology research articles can be understood and used by everyone

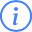
 Note

If you have not used a reporting guideline before, read about [how and why to use them](https:/resources.equator-network.org/about/reporting-guidelines.html) and check whether STROBE is the [most applicable reporting guideline](https:/resources.equator-network.org/reporting-guidelines/strobe/index.html?#applicability) for your work.

Reporting guidelines are most useful when used early in research. When writing a manuscript or application, consider using the [Full Guidance](https:/resources.equator-network.org/reporting-guidelines/strobe/index.html) where you’ll see explanations and examples for each item.

After writing, demonstrate adherence by completing this checklist:

1. Specify where each item is described (see Note 1).
2. Cite this checklist (See Note 2).
3. Include your completed checklist as a supplement when submitting to a journal so that future readers can use it to find information.

|  | Item Description | Location (or reason for not reporting) |
| --- | --- | --- |
| **Title and abstract** |  |  |
| [1a. Indicate the study’s design](https:/resources.equator-network.org/reporting-guidelines/strobe/items/title-abstract-indicate-study-design.html?utm_source=strobe&utm_medium=checklist&utm_campaign=1_1) | Indicate the study’s design with a commonly used term in the title or the abstract. | Title and Abstract |
| [1b. Abstract](https:/resources.equator-network.org/reporting-guidelines/strobe/items/abstract.html?utm_source=strobe&utm_medium=checklist&utm_campaign=1_1) | Provide in the abstract an informative and balanced summary of what was done and what was found. | Background , Methods, Results, Conclusion |
| **Introduction** |  |  |
| [2. Background / rationale](https:/resources.equator-network.org/reporting-guidelines/strobe/items/background-rationale.html?utm_source=strobe&utm_medium=checklist&utm_campaign=1_1) | Explain the scientific background and rationale for the investigation being reported. | Row 103-112 |
| [3. Objectives](https:/resources.equator-network.org/reporting-guidelines/strobe/items/objectives.html?utm_source=strobe&utm_medium=checklist&utm_campaign=1_1) | State specific objectives, including any prespecified hypotheses. | Introduction – final paragraph |
| **Methods** |  |  |
| [4. Study design](https:/resources.equator-network.org/reporting-guidelines/strobe/items/study-design.html?utm_source=strobe&utm_medium=checklist&utm_campaign=1_1) | Present key elements of study design early in the paper. | Methods  (Observational study integrating clinical follow-up with computational modeling) |
| [5. Setting](https:/resources.equator-network.org/reporting-guidelines/strobe/items/setting.html?utm_source=strobe&utm_medium=checklist&utm_campaign=1_1) | Describe the setting, locations, and relevant dates, including periods of recruitment, exposure, follow-up, and data collection. | Methods – Clinical evaluation section  (Pediatric neurology setting; follow-up duration described) |
| [6a. Eligibility criteria](https:/resources.equator-network.org/reporting-guidelines/strobe/items/eligibility-criteria.html?utm_source=strobe&utm_medium=checklist&utm_campaign=1_1) | **Cohort study:** Give the eligibility criteria, and the sources and methods of selection of participants. Describe methods of follow-up. **Case-control study:** Give the eligibility criteria, and the sources and methods of case ascertainment and control selection. Give the rationale for the choice of cases and controls. **Cross-sectional study:** Give the eligibility criteria, and the sources and methods of selection of participants. | Methods – Clinical evaluation  (Genetically confirmed *ADPRHL2*-related neurodegeneration) |
| [6b. Matching criteria](https:/resources.equator-network.org/reporting-guidelines/strobe/items/matching-criteria.html?utm_source=strobe&utm_medium=checklist&utm_campaign=1_1) | **Cohort study:** For matched studies, give matching criteria and number of exposed and unexposed. **Case-control study:** For matched studies, give matching criteria and the number of controls per case. | Not applicable  (Single observational translational analysis; no comparison groups) |
| [7. Variables](https:/resources.equator-network.org/reporting-guidelines/strobe/items/variables.html?utm_source=strobe&utm_medium=checklist&utm_campaign=1_1) | Clearly define all outcomes, exposures, predictors, potential confounders, and effect modifiers. Give diagnostic criteria, if applicable. | Methods – Clinical assessment and In-silico analysis sections  (Neurological status, motor function, protein stability metrics) |
| [8. Data sources / measurement](https:/resources.equator-network.org/reporting-guidelines/strobe/items/data-sources-measurement.html?utm_source=strobe&utm_medium=checklist&utm_campaign=1_1) | For each variable of interest give sources of data and details of methods of assessment (measurement). Describe comparability of assessment methods if there is more than one group. | Methods – Clinical evaluation; Molecular modeling and MD simulatio |
| [9. Bias](https:/resources.equator-network.org/reporting-guidelines/strobe/items/bias.html?utm_source=strobe&utm_medium=checklist&utm_campaign=1_1) | Describe any efforts to address potential sources of bias. | Discussion – Limitations paragraph  (Potential confounding by developmental trajectory acknowledged) |
| [10. Study size](https:/resources.equator-network.org/reporting-guidelines/strobe/items/study-size.html?utm_source=strobe&utm_medium=checklist&utm_campaign=1_1) | Explain how the study size was arrived at. | (Study size determined by availability of genetically confirmed case due to ultra-rare disease) |
| [11. Quantitative variables](https:/resources.equator-network.org/reporting-guidelines/strobe/items/quantitative-variables.html?utm_source=strobe&utm_medium=checklist&utm_campaign=1_1) | Explain how quantitative variables were handled in the analyses. If applicable, describe which groupings were chosen, and why. | Methods – Molecular dynamics analysis |
| [12a. Statistical methods](https:/resources.equator-network.org/reporting-guidelines/strobe/items/statistical-methods-description.html?utm_source=strobe&utm_medium=checklist&utm_campaign=1_1) | Describe all statistical methods, including those used to control for confounding. | Methods – In-silico analysis |
| [12b. Statistical methods – subgroups and interactions](https:/resources.equator-network.org/reporting-guidelines/strobe/items/statistical-methods-subgroups-interactions.html?utm_source=strobe&utm_medium=checklist&utm_campaign=1_1) | Describe any methods used to examine subgroups and interactions. | Not applicable |
| [12c. Statistical methods – missing data](https:/resources.equator-network.org/reporting-guidelines/strobe/items/statistical-methods-missing-data.html?utm_source=strobe&utm_medium=checklist&utm_campaign=1_1) | Explain how missing data were addressed. | No missing computational or clinical follow-up data |
| [12di. Statistical methods – loss to follow-up](https:/resources.equator-network.org/reporting-guidelines/strobe/items/statistical-methods-loss-to-follow-up.html?utm_source=strobe&utm_medium=checklist&utm_campaign=1_1) | **Cohort study:** If applicable, describe how loss to follow-up was addressed. | Not applicable |
| [12dii. Statistical methods – matching cases and controls](https:/resources.equator-network.org/reporting-guidelines/strobe/items/statistical-methods-matching-cases-controls.html?utm_source=strobe&utm_medium=checklist&utm_campaign=1_1) | **Case-control study:** If applicable, explain how matching of cases and controls was addressed. | Not applicable |
| [12diii. Statistical methods – sampling strategy](https:/resources.equator-network.org/reporting-guidelines/strobe/items/statistical-methods-analytical-methods-sampling-strategy.html?utm_source=strobe&utm_medium=checklist&utm_campaign=1_1) | **Cross-sectional study:** If applicable, describe analytical methods taking account of sampling strategy. | Not applicable |
| [12e. Statistical methods – sensitivity analyses](https:/resources.equator-network.org/reporting-guidelines/strobe/items/statistical-methods-sensitivity-analyses.html?utm_source=strobe&utm_medium=checklist&utm_campaign=1_1) | Describe any sensitivity analyses. | Results – Molecular dynamics analysis |
| **Results** |  |  |
| [13a. Participant numbers](https:/resources.equator-network.org/reporting-guidelines/strobe/items/participants-numbers.html?utm_source=strobe&utm_medium=checklist&utm_campaign=1_1) | Report the numbers of individuals at each stage of the study—e.g., numbers potentially eligible, examined for eligibility, confirmed eligible, included in the study, completing follow-up, and analysed; Consider use of a flow diagram. | Methods – Clinical evaluation |
| [13b. Participants – non-participation](https:/resources.equator-network.org/reporting-guidelines/strobe/items/participants-non-participation.html?utm_source=strobe&utm_medium=checklist&utm_campaign=1_1) | Give reasons for non-participation at each stage. | Not applicable |
| [13c. Participants – flow diagram](https:/resources.equator-network.org/reporting-guidelines/strobe/items/participants-flow-diagram.html?utm_source=strobe&utm_medium=checklist&utm_campaign=1_1) | Consider use of a flow diagram. | Not applicable |
| [14a. Descriptive data – participant characteristics](https:/resources.equator-network.org/reporting-guidelines/strobe/items/descriptive-data-participant-characteristics.html?utm_source=strobe&utm_medium=checklist&utm_campaign=1_1) | Give characteristics of study participants (e.g., demographic, clinical, social) and information on exposures and potential confounders. Present the information in a table. | Results – Clinical findings |
| [14b. Descriptive data – missing data](https:/resources.equator-network.org/reporting-guidelines/strobe/items/descriptive-data-missing-data.html?utm_source=strobe&utm_medium=checklist&utm_campaign=1_1) | Indicate the number of participants with missing data for each variable of interest. | Not applicable |
| [14c. Descriptive data – follow-up time](https:/resources.equator-network.org/reporting-guidelines/strobe/items/descriptive-data-follow-up-time.html?utm_source=strobe&utm_medium=checklist&utm_campaign=1_1) | **Cohort study:** Summarise follow-up time—e.g., average and total amount. | Clinical follow-up (12 months) |
| [15. Outcome data](https:/resources.equator-network.org/reporting-guidelines/strobe/items/outcome-data.html?utm_source=strobe&utm_medium=checklist&utm_campaign=1_1) | **Cohort study:** Report numbers of outcome events or summary measures over time. **Case-control study:** Report numbers in each exposure category, or summary measures of exposure. **Cross-sectional study:** Report numbers of outcome events or summary measures. | Clinical outcomes; MD simulation results |
| [16a. Main results](https:/resources.equator-network.org/reporting-guidelines/strobe/items/main-results.html?utm_source=strobe&utm_medium=checklist&utm_campaign=1_1) | Give unadjusted estimates and, if applicable, confounder-adjusted estimates and their precision (e.g., 95% confidence intervals). Make clear which confounders were adjusted for and why they were included. | Section 3 |
| [16b. Main results – category boundaries](https:/resources.equator-network.org/reporting-guidelines/strobe/items/main-results-category-boundaries.html?utm_source=strobe&utm_medium=checklist&utm_campaign=1_1) | Report category boundaries when continuous variables were categorised. | Not applicable |
| [16c. Main results – risk](https:/resources.equator-network.org/reporting-guidelines/strobe/items/main-results-risk.html?utm_source=strobe&utm_medium=checklist&utm_campaign=1_1) | If relevant, consider translating estimates of relative risk into absolute risk for a meaningful time period. | Not applicable |
| [17. Other analyses](https:/resources.equator-network.org/reporting-guidelines/strobe/items/other-analyses.html?utm_source=strobe&utm_medium=checklist&utm_campaign=1_1) | Report other analyses done—e.g., analyses of subgroups and interactions, and sensitivity analyses. | Molecular dynamics analyses |
| **Discussion** |  |  |
| [18. Key results](https:/resources.equator-network.org/reporting-guidelines/strobe/items/key-results.html?utm_source=strobe&utm_medium=checklist&utm_campaign=1_1) | Summarise key results with reference to study objectives. | 308-332 |
| [19. Limitations](https:/resources.equator-network.org/reporting-guidelines/strobe/items/limitations.html?utm_source=strobe&utm_medium=checklist&utm_campaign=1_1) | Discuss limitations of the study, taking into account sources of potential bias or imprecision. Discuss both direction and magnitude of any potential bias. | Row 340-348 |
| [20. Interpretation](https:/resources.equator-network.org/reporting-guidelines/strobe/items/interpretation.html?utm_source=strobe&utm_medium=checklist&utm_campaign=1_1) | Give a cautious overall interpretation considering objectives, limitations, multiplicity of analyses, results from similar studies, and other relevant evidence. | row 334-348 |
| [21. Generalisability](https:/resources.equator-network.org/reporting-guidelines/strobe/items/generalisability.html?utm_source=strobe&utm_medium=checklist&utm_campaign=1_1) | Discuss the generalisability (external validity) of the study results. | row 334-340 |
| **Other information** |  |  |
| [22. Funding](https:/resources.equator-network.org/reporting-guidelines/strobe/items/funding.html?utm_source=strobe&utm_medium=checklist&utm_campaign=1_1) | Give the source of funding and the role of the funders for the present study and, if applicable, for the original study on which the present article is based. | Row389 |

### 1 How to specify where content is

Tell the reader where they can find information. E.g.,

- Results; paragraph 2
- Methods, Participants; paragraphs 1 & 2.
- Table 3
- Supplement B, para. 4

If you have chosen not to describe an item, explain why. You can do this in the checklist, or as a note below it.

You can describe items in the article body, or in tables, figures, or supplementary materials, and should prioritize items you feel are most important to your intended audience. The order of items in your manuscript does not need to match the order of items in this checklist. You can decide how best to structure your work.

### 2 How to cite

Describe how you used STROBE at the end of your Methods section, referencing the resources you used e.g.,

‘We used the STROBE reporting guideline(1) to draft this manuscript, and the STROBE reporting checklist(2) when editing, included in supplement A’

If you use a reporting checklist, remember to include it as a supplement when publishing so that readers can easily find information and see how you have interpreted the guidance.

1.	Elm E von, Altman DG, Egger M, Pocock SJ, Gøtzsche PC, Vandenbroucke JP, et al. The strengthening the reporting of observational studies in epidemiology (STROBE) statement: Guidelines for reporting observational studies. Annals of Internal Medicine [Internet]. 2007 Oct;147(8):573–7. Available from: <https://www.acpjournals.org/doi/10.7326/0003-4819-147-8-200710160-00010>

2.	Elm E von, Altman DG, Egger M, Pocock SJ, Gøtzsche PC, Vandenbroucke JP, et al. The STROBE reporting checklist. In: Harwood J, Albury C, Beyer J de, Schlüssel M, Collins G, editors. The EQUATOR network reporting guideline platform [Internet]. The UK EQUATOR Centre; 2025. Available from: [https:/resources.equator-network.org/reporting-guidelines/strobe/strobe-checklist.docx](https://https:/resources.equator-network.org/reporting-guidelines/strobe/strobe-checklist.docx)
